## Supplementary material for "Racial and ethnic differentials in COVID-19-related job exposures by occupational standing in the US": S1 Appendix

**S1 Appendix: Racial/ethnic distributions within each occupational subset displayed in Main Text Figures**

*Updated April 5, 2021*

**Table S1.1: Racial/ethnic distribution of all recent workers by sex**

| Race/Ethnicity | Male | | Female | |
|---|---|---|---|---|
| | N | Percent (%) | N | Percent (%) |
| White | 675,542 | 63.1 | 632,421 | 62.4 |
| Black | 76,649 | 10.6 | 87,168 | 12.7 |
| Latino | 133,868 | 17.7 | 120,853 | 15.8 |
| Chinese | 13,095 | 1.3 | 14,265 | 1.5 |
| Filipino | 7,798 | 0.8 | 10,403 | 1.1 |
| Vietnamese | 5,150 | 0.6 | 5,519 | 0.6 |
| Korean | 4,303 | 0.5 | 4,684 | 0.5 |
| Other Asian | 22,072 | 2.5 | 18,654 | 2.2 |
| Native American | 7,018 | 0.6 | 7,059 | 0.6 |
| Pacific Islander | 1,402 | 0.2 | 1,353 | 0.2 |
| Mixed Race | 17,555 | 2.0 | 17,857 | 2.1 |
| Other Race | 1,695 | 0.2 | 1,778 | 0.2 |
| Total | 966,147 | 100 | 922,014 | 100 |

Note: Data are from the 2018 American Community Survey (ACS). Percentages are calculated using weights provided by the ACS; sample sizes (N) are unweighted. The percentages in this table correspond to those displayed in panel 1 in Figs 1-2.

**Table S1.2: Racial/ethnic distribution of male frontline workers by quartile of occupational standing**

| Race/Ethnicity | Quartile of occupational standing | | | | | |
|---|---|---|---|---|---|---|
| | 1st Quartile | 2nd Quartile | 3rd Quartile | 4th Quartile | All frontline workers | Total N |
| White | 53.6 | 62.1 | 71.2 | 69.6 | 59.0 | 399,608 |
| Black | 13.5 | 12.4 | 8.4 | 6.6 | 12.2 | 55,605 |
| Latino | 26.6 | 17.8 | 12.9 | 8.5 | 21.6 | 101,260 |
| Chinese | 0.7 | 0.9 | 1.0 | 2.5 | 0.9 | 5,257 |
| Filipino | 0.6 | 0.9 | 0.8 | 2.8 | 0.8 | 4,751 |
| Vietnamese | 0.5 | 0.8 | 0.4 | 1.0 | 0.6 | 3,236 |
| Korean | 0.2 | 0.4 | 0.5 | 1.1 | 0.3 | 1,881 |
| Other Asian | 1.3 | 1.7 | 2.1 | 5.3 | 1.7 | 9,240 |
| Native American | 0.8 | 0.6 | 0.5 | 0.3 | 0.7 | 5,329 |
| Pacific Islander | 0.2 | 0.2 | 0.1 | 0.1 | 0.2 | 1,009 |
| Mixed Race | 1.9 | 2.0 | 2.0 | 2.1 | 1.9 | 11,022 |
| Other Race | 0.2 | 0.2 | 0.2 | 0.3 | 0.2 | 1,077 |
| Total Percent | 100 | 100 | 100 | 100 | 100 | |
| Total N | 327,280 | 154,040 | 91,625 | 26,330 | 599,275 | |

Note: Data are from the 2018 American Community Survey (ACS). Percentages are calculated using weights provided by the ACS; sample sizes (N) are unweighted. Occupational standing is defined as the percentage of ACS respondents reporting this occupation title who completed at least one year of college education. Occupational standing is then divided into quartiles. The 1st quartile represents the lowest quartile of occupational standing and the 4th quartile the highest quartile of occupational standing. The percentages in this table correspond to those displayed in panel 2 of Figs 1 and 3.

**Table S1.3: Racial/ethnic distribution of female frontline workers by quartile of occupational standing**

| Race/Ethnicity | Quartile of occupational standing | | | | | |
|---|---|---|---|---|---|---|
| | 1st Quartile | 2nd Quartile | 3rd Quartile | 4th Quartile | All frontline workers | Total N |
| White | 47.6 | 55.2 | 65.8 | 71.7 | 57.0 | 285,517 |
| Black | 15.7 | 17.0 | 11.8 | 9.4 | 14.5 | 48,456 |
| Latino | 28.3 | 18.1 | 14.3 | 7.2 | 19.3 | 70,995 |
| Chinese | 1.1 | 1.3 | 1.2 | 1.5 | 1.2 | 5,765 |
| Filipino | 1.0 | 1.3 | 1.1 | 3.0 | 1.4 | 6,232 |
| Vietnamese | 0.7 | 1.3 | 0.4 | 0.7 | 0.8 | 3,542 |
| Korean | 0.4 | 0.5 | 0.5 | 0.8 | 0.5 | 2,189 |
| Other Asian | 1.9 | 1.8 | 1.6 | 3.3 | 2.0 | 8,247 |
| Native American | 0.9 | 0.6 | 0.6 | 0.3 | 0.7 | 3,989 |
| Pacific Islander | 0.2 | 0.2 | 0.2 | 0.1 | 0.2 | 719 |
| Mixed Race | 2.1 | 2.3 | 2.2 | 1.9 | 2.2 | 8,948 |
| Other Race | 0.3 | 0.3 | 0.2 | 0.2 | 0.3 | 963 |
| Total Percent | 100 | 100 | 100 | 100 | 100 | |
| Total N | 146,643 | 140,283 | 92,265 | 66,371 | 445,562 | |

Note: Data are from the 2018 American Community Survey (ACS). Percentages are calculated using weights provided by the ACS; sample sizes (N) are unweighted. Occupational standing is defined as the percentage of ACS respondents reporting this occupation title who completed at least one year of college education. Occupational standing is then divided into quartiles. The 1st quartile represents the lowest quartile of occupational standing and the 4th quartile the highest quartile of occupational standing. The percentages in this table correspond to those displayed in panel 2 of Figs 2 and 4.

**Table S1.4: Racial/ethnic distribution of male frontline workers at highest risk of exposure to infection by quartile of occupational standing**

| Race/Ethnicity | Quartile of occupational standing | | | | | |
|---|---|---|---|---|---|---|
| | 1st Quartile | 2nd Quartile | 3rd Quartile | 4th Quartile | All in risk quartile | Total N |
| White | 51.7 | 57.4 | 65.4 | 67.2 | 59.1 | 71,144 |
| Black | 17.1 | 17.7 | 12.5 | 7.3 | 14.4 | 11,717 |
| Latino | 24.8 | 16.9 | 14.4 | 8.5 | 17.4 | 15,040 |
| Chinese | 0.6 | 0.9 | 0.8 | 2.7 | 1.1 | 1,234 |
| Filipino | 0.9 | 1.6 | 1.4 | 3.3 | 1.6 | 1,728 |
| Vietnamese | 0.4 | 0.7 | 0.4 | 1.1 | 0.6 | 628 |
| Korean | 0.3 | 0.4 | 0.2 | 1.1 | 0.5 | 487 |
| Other Asian | 1.0 | 1.4 | 1.5 | 6.0 | 2.2 | 2,187 |
| Native American | 1.1 | 0.7 | 0.7 | 0.3 | 0.7 | 1,120 |
| Pacific Islander | 0.2 | 0.2 | 0.2 | 0.1 | 0.2 | 190 |
| Mixed Race | 1.9 | 1.9 | 2.4 | 2.0 | 2.0 | 2,060 |
| Other Race | 0.2 | 0.3 | 0.3 | 0.3 | 0.2 | 200 |
| Total Percent | 100 | 100 | 100 | 100 | 100 | |
| Total N | 34,776 | 27,899 | 23,147 | 21,913 | 107,735 | |

Note: Data are from the 2018 American Community Survey (ACS). Percentages are calculated using weights provided by the ACS; sample sizes (N) are unweighted. Workers are considered to be at highest risk of exposure to infection if the average exposure to infection for their occupation in the O*NET data falls in the highest quartile of the occupations in the analysis. Occupational standing is defined as the percentage of ACS respondents reporting this occupation title who completed at least one year of college education. Occupational standing is then divided into quartiles. The 1st quartile represents the lowest quartile of occupational standing and the 4th quartile the highest quartile of occupational standing. The percentages in this table correspond to those displayed in panel 3 of Figs 1 and 3.

**Table S1.5: Racial/ethnic distribution of male frontline workers at highest risk of physical proximity to others by quartile of occupational standing**

| Race/Ethnicity | Quartile of occupational standing | | | | | |
|---|---|---|---|---|---|---|
| | 1st Quartile | 2nd Quartile | 3rd Quartile | 4th Quartile | All in risk quartile | Total N |
| White | 53.5 | 55.9 | 64.1 | 69.5 | 58.7 | 77,439 |
| Black | 9.0 | 14.9 | 12.3 | 6.5 | 11.6 | 10,374 |
| Latino | 31.9 | 19.1 | 14.5 | 8.4 | 20.0 | 18,645 |
| Chinese | 0.6 | 1.4 | 1.0 | 2.4 | 1.3 | 1,597 |
| Filipino | 0.6 | 1.3 | 1.7 | 3.1 | 1.5 | 1,711 |
| Vietnamese | 0.3 | 1.4 | 0.5 | 0.8 | 0.9 | 948 |
| Korean | 0.2 | 0.5 | 0.3 | 1.0 | 0.5 | 547 |
| Other Asian | 0.8 | 2.4 | 1.8 | 5.7 | 2.5 | 2,738 |
| Native American | 0.8 | 0.6 | 0.6 | 0.3 | 0.6 | 988 |
| Pacific Islander | 0.2 | 0.2 | 0.2 | 0.1 | 0.2 | 181 |
| Mixed Race | 1.9 | 2.1 | 2.7 | 2.1 | 2.1 | 2,427 |
| Other Race | 0.2 | 0.3 | 0.3 | 0.3 | 0.3 | 242 |
| Total Percent | 100 | 100 | 100 | 100 | 100 | |
| Total N | 28,853 | 50,772 | 17,301 | 20,911 | 11,7837 | |

Note: Data are from the 2018 American Community Survey (ACS). Percentages are calculated using weights provided by the ACS; sample sizes (N) are unweighted. Workers are considered to be at highest risk of physical proximity to others if the average level of physical proximity to others for their occupation in the O*NET data falls in the highest quartile of the occupations in the analysis. Occupational standing is defined as the percentage of ACS respondents reporting this occupation title who completed at least one year of college education. Occupational standing is then divided into quartiles. The 1st quartile represents the lowest quartile of occupational standing and the 4th quartile the highest quartile of occupational standing. The percentages in this table correspond to those displayed in panel 4 of Figs 1 and 3.

**Table S1.6: Racial/ethnic distribution of male frontline workers at highest risk of face-to-face discussions by quartile of occupational standing**

| Race/Ethnicity | Quartile of occupational standing | | | | | |
|---|---|---|---|---|---|---|
| | 1st Quartile | 2nd Quartile | 3rd Quartile | 4th Quartile | All in risk quartile | Total N |
| White | 56.0 | 67.4 | 70.6 | 68.3 | 64.1 | 111,245 |
| Black | 14.8 | 9.3 | 8.2 | 6.6 | 10.7 | 12,335 |
| Latino | 23.4 | 17.2 | 13.0 | 7.6 | 17.4 | 21,536 |
| Chinese | 0.4 | 0.7 | 1.0 | 2.9 | 0.9 | 1,460 |
| Filipino | 0.6 | 0.6 | 0.8 | 3.3 | 0.9 | 1,432 |
| Vietnamese | 0.3 | 0.4 | 0.4 | 1.2 | 0.5 | 689 |
| Korean | 0.2 | 0.3 | 0.7 | 1.1 | 0.4 | 659 |
| Other Asian | 1.0 | 1.3 | 2.8 | 6.5 | 2.1 | 2,996 |
| Native American | 0.7 | 0.6 | 0.5 | 0.2 | 0.6 | 1,104 |
| Pacific Islander | 0.2 | 0.1 | 0.1 | 0.1 | 0.2 | 238 |
| Mixed Race | 2.2 | 1.8 | 1.7 | 2.0 | 2.0 | 2,826 |
| Other Race | 0.2 | 0.2 | 0.2 | 0.3 | 0.2 | 265 |
| Total Percent | 100 | 100 | 100 | 100 | 100 | |
| Total N | 54,074 | 48,152 | 36,232 | 18,327 | 156,785 | |

Note: Data are from the 2018 American Community Survey (ACS). Percentages are calculated using weights provided by the ACS; sample sizes (N) are unweighted. Workers are considered to be at highest risk of face-to-face discussions if the average level of face-to-face discussions for their occupation in the O*NET data falls in the highest quartile of the occupations in the analysis. Occupational standing is defined as the percentage of ACS respondents reporting this occupation title who completed at least one year of college education. Occupational standing is then divided into quartiles. The 1st quartile represents the lowest quartile of occupational standing and the 4th quartile the highest quartile of occupational standing. The percentages in this table correspond to those displayed in panel 5 of Figs 1 and 3.

**Table S1.7: Racial/ethnic distribution of male frontline workers at highest risk of interactions with external customers or the public by quartile of occupational standing**

| Race/Ethnicity | Quartile of occupational standing | | | | | |
|---|---|---|---|---|---|---|
| | 1st Quartile | 2nd Quartile | 3rd Quartile | 4th Quartile | All in risk quartile | Total N |
| White | 51.8 | 63.1 | 73.2 | 71.2 | 67.5 | 96,530 |
| Black | 14.4 | 11.5 | 7.2 | 5.6 | 9.3 | 8,933 |
| Latino | 21.6 | 17.7 | 12.6 | 8.0 | 15.0 | 16,183 |
| Chinese | 1.2 | 0.9 | 0.9 | 3.0 | 1.0 | 1,449 |
| Filipino | 1.0 | 0.9 | 0.6 | 2.1 | 0.8 | 985 |
| Vietnamese | 0.7 | 0.5 | 0.3 | 1.7 | 0.5 | 613 |
| Korean | 0.6 | 0.4 | 0.5 | 1.3 | 0.5 | 660 |
| Other Asian | 4.3 | 1.6 | 2.0 | 4.8 | 2.3 | 2,710 |
| Native American | 0.8 | 0.6 | 0.5 | 0.1 | 0.5 | 825 |
| Pacific Islander | 0.1 | 0.1 | 0.1 | 0.1 | 0.1 | 166 |
| Mixed Race | 3.3 | 2.4 | 1.9 | 1.8 | 2.2 | 2,700 |
| Other Race | 0.2 | 0.3 | 0.2 | 0.2 | 0.2 | 227 |
| Total Percent | 100 | 100 | 100 | 100 | 100 | |
| Total N | 14,600 | 39,186 | 71,092 | 7,103 | 131,981 | |

Note: Data are from the 2018 American Community Survey (ACS). Percentages are calculated using weights provided by the ACS; sample sizes (N) are unweighted. Workers are considered to be at highest risk of interactions with external customers or the public if the average level of interactions with external customers or the public for their occupation in the O*NET data falls in the highest quartile of the occupations in the analysis. Occupational standing is defined as the percentage of ACS respondents reporting this occupation title who completed at least one year of college education. Occupational standing is then divided into quartiles. The 1st quartile represents the lowest quartile of occupational standing and the 4th quartile the highest quartile of occupational standing. The percentages in this table correspond to those displayed in panel 6 of Figs 1 and 3.

**Table S1.8: Racial/ethnic distribution of male frontline workers at highest risk of working indoors by quartile of occupational standing**

| Race/Ethnicity | Quartile of occupational standing | | | | | |
|---|---|---|---|---|---|---|
| | 1st Quartile | 2nd Quartile | 3rd Quartile | 4th Quartile | All in risk quartile | Total N |
| White | 50.8 | 52.6 | 58.2 | 67.4 | 62.2 | 22,561 |
| Black | 15.8 | 11.2 | 14.1 | 7.1 | 9.8 | 2,371 |
| Latino | 27.2 | 21.8 | 16.2 | 8.5 | 13.7 | 3,638 |
| Chinese | 0.9 | 2.7 | 1.4 | 3.0 | 2.4 | 813 |
| Filipino | 0.5 | 1.5 | 3.2 | 3.0 | 2.5 | 796 |
| Vietnamese | 1.1 | 2.0 | 0.6 | 1.2 | 1.2 | 373 |
| Korean | 0.4 | 1.2 | 0.3 | 1.2 | 1.0 | 328 |
| Other Asian | 1.1 | 3.3 | 2.1 | 6.1 | 4.5 | 1,409 |
| Native American | 0.7 | 1.0 | 0.6 | 0.3 | 0.5 | 185 |
| Pacific Islander | 0.1 | 0.5 | 0.2 | 0.1 | 0.2 | 44 |
| Mixed Race | 1.3 | 2.0 | 2.7 | 1.9 | 2.0 | 628 |
| Other Race | 0.2 | 0.3 | 0.4 | 0.3 | 0.3 | 67 |
| Total Percent | 100 | 100 | 100 | 100 | 100 | |
| Total N | 4,281 | 3,596 | 4,332 | 21,004 | 33,213 | |

Note: Data are from the 2018 American Community Survey (ACS). Percentages are calculated using weights provided by the ACS; sample sizes (N) are unweighted. Because working indoors rather than outdoors reflects high risk, we take a maximum of the O*NET values for two variables measuring working outdoors and designate workers whose occupations fall in the lowest quartile of this measure as having the highest risk of working indoors. Occupational standing is defined as the percentage of ACS respondents reporting this occupation title who completed at least one year of college education. Occupational standing is then divided into quartiles. The 1st quartile represents the lowest quartile of occupational standing and the 4th quartile the highest quartile of occupational standing. The percentages in this table correspond to those displayed in panel 7 of Figs 1 and 3.

**Table S1.9: Racial/ethnic distribution of female frontline workers at highest risk of exposure to infection by quartile of occupational standing**

| Race/Ethnicity | Quartile of occupational standing | | | | | |
|---|---|---|---|---|---|---|
| | 1st Quartile | 2nd Quartile | 3rd Quartile | 4th Quartile | All in risk quartile | Total N |
| White | 38.0 | 52.7 | 59.2 | 71.8 | 57.3 | 129,213 |
| Black | 15.3 | 21.3 | 16.2 | 9.5 | 15.8 | 23,792 |
| Latino | 40.2 | 18.0 | 16.4 | 7.1 | 18.0 | 29,777 |
| Chinese | 0.8 | 1.1 | 1.0 | 1.5 | 1.1 | 2,379 |
| Filipino | 1.1 | 1.4 | 1.6 | 3.0 | 1.9 | 3,819 |
| Vietnamese | 0.4 | 0.5 | 0.4 | 0.7 | 0.5 | 1,053 |
| Korean | 0.2 | 0.3 | 0.4 | 0.8 | 0.5 | 959 |
| Other Asian | 1.1 | 1.5 | 1.6 | 3.2 | 2.0 | 3,703 |
| Native American | 1.0 | 0.7 | 0.7 | 0.3 | 0.6 | 1,726 |
| Pacific Islander | 0.2 | 0.2 | 0.1 | 0.1 | 0.2 | 290 |
| Mixed Race | 1.3 | 2.0 | 2.1 | 1.8 | 1.9 | 3,514 |
| Other Race | 0.5 | 0.3 | 0.3 | 0.2 | 0.3 | 459 |
| Total Percent | 100 | 100 | 100 | 100 | 100 | |
| Total N | 29,869 | 68,645 | 36,755 | 65,415 | 200,684 | |

Note: Data are from the 2018 American Community Survey (ACS). Percentages are calculated using weights provided by the ACS; sample sizes (N) are unweighted. Workers are considered to be at highest risk of exposure to infection if the average exposure to infection for their occupation in the O*NET data falls in the highest quartile of the occupations in the analysis. Occupational standing is defined as the percentage of ACS respondents reporting this occupation title who completed at least one year of college education. Occupational standing is then divided into quartiles. The 1st quartile represents the lowest quartile of occupational standing and the 4th quartile the highest quartile of occupational standing. The percentages in this table correspond to those displayed in panel 3 of Figs 2 and 4.

**Table S1.10: Racial/ethnic distribution of female frontline workers at highest risk of physical proximity to others by quartile of occupational standing**

| Race/Ethnicity | Quartile of occupational standing | | | | | |
|---|---|---|---|---|---|---|
| | 1st Quartile | 2nd Quartile | 3rd Quartile | 4th Quartile | All in risk quartile | Total N |
| White | 56.2 | 53.2 | 62.0 | 72.6 | 60.2 | 137,670 |
| Black | 12.7 | 18.5 | 14.4 | 9.2 | 14.9 | 22,852 |
| Latino | 22.6 | 18.1 | 15.6 | 7.0 | 15.0 | 26,002 |
| Chinese | 1.2 | 1.5 | 0.9 | 1.4 | 1.3 | 2,786 |
| Filipino | 0.9 | 1.4 | 1.5 | 3.1 | 1.8 | 3,768 |
| Vietnamese | 0.6 | 1.7 | 0.4 | 0.5 | 1.1 | 2,094 |
| Korean | 0.4 | 0.5 | 0.4 | 0.7 | 0.5 | 1,041 |
| Other Asian | 1.5 | 1.8 | 1.5 | 3.0 | 2.0 | 3,973 |
| Native American | 0.6 | 0.7 | 0.7 | 0.3 | 0.6 | 1,663 |
| Pacific Islander | 0.3 | 0.2 | 0.1 | 0.1 | 0.2 | 289 |
| Mixed Race | 2.9 | 2.3 | 2.3 | 1.8 | 2.2 | 4,119 |
| Other Race | 0.2 | 0.3 | 0.2 | 0.2 | 0.3 | 438 |
| Total Percent | 100 | 100 | 100 | 100 | 100 | |
| Total N | 12,673 | 93,583 | 41,868 | 58,571 | 206,695 | |

Note: Data are from the 2018 American Community Survey (ACS). Percentages are calculated using weights provided by the ACS; sample sizes (N) are unweighted. Workers are considered to be at highest risk of physical proximity to others if the average level of physical proximity to others for their occupation in the O*NET data falls in the highest quartile of the occupations in the analysis. Occupational standing is defined as the percentage of ACS respondents reporting this occupation title who completed at least one year of college education. Occupational standing is then divided into quartiles. The 1st quartile represents the lowest quartile of occupational standing and the 4th quartile the highest quartile of occupational standing. The percentages in this table correspond to those displayed in panel 4 of Figs 2 and 4.

**Table S1.11: Racial/ethnic distribution of female frontline workers at highest risk of face-to-face discussions by quartile of occupational standing**

| Race/Ethnicity | Quartile of occupational standing | | | | | |
|---|---|---|---|---|---|---|
| | 1st Quartile | 2nd Quartile | 3rd Quartile | 4th Quartile | All in risk quartile | Total N |
| White | 53.8 | 63.2 | 64.2 | 71.3 | 65.4 | 102,688 |
| Black | 16.6 | 11.5 | 14.0 | 9.6 | 12.0 | 12,479 |
| Latino | 21.6 | 16.9 | 13.8 | 6.9 | 12.9 | 15,405 |
| Chinese | 0.7 | 0.9 | 1.0 | 1.5 | 1.2 | 1,728 |
| Filipino | 0.9 | 1.0 | 1.2 | 3.2 | 1.9 | 2,633 |
| Vietnamese | 0.5 | 0.8 | 0.3 | 0.7 | 0.6 | 818 |
| Korean | 0.3 | 0.5 | 0.6 | 0.9 | 0.7 | 903 |
| Other Asian | 1.9 | 1.9 | 1.7 | 3.4 | 2.4 | 3,116 |
| Native American | 0.9 | 0.5 | 0.7 | 0.3 | 0.5 | 819 |
| Pacific Islander | 0.2 | 0.2 | 0.2 | 0.1 | 0.1 | 176 |
| Mixed Race | 2.3 | 2.4 | 2.1 | 1.9 | 2.1 | 2,660 |
| Other Race | 0.3 | 0.3 | 0.2 | 0.2 | 0.2 | 263 |
| Total Percent | 100 | 100 | 100 | 100 | 100 | |
| Total N | 14,683 | 36,851 | 36,626 | 55,528 | 143,688 | |

Note: Data are from the 2018 American Community Survey (ACS). Percentages are calculated using weights provided by the ACS; sample sizes (N) are unweighted. Workers are considered to be at highest risk of face-to-face discussions if the average level of face-to-face discussions for their occupation in the O*NET data falls in the highest quartile of the occupations in the analysis. Occupational standing is defined as the percentage of ACS respondents reporting this occupation title who completed at least one year of college education. Occupational standing is then divided into quartiles. The 1st quartile represents the lowest quartile of occupational standing and the 4th quartile the highest quartile of occupational standing. The percentages in this table correspond to those displayed in panel 5 of Figs 2 and 4.

**Table S1.12: Racial/ethnic distribution of female frontline workers at highest risk of interactions with external customers or the public by quartile of occupational standing**

| Race/Ethnicity | Quartile of occupational standing | | | | | |
|---|---|---|---|---|---|---|
| | 1st Quartile | 2nd Quartile | 3rd Quartile | 4th Quartile | All in risk quartile | Total N |
| White | 51.9 | 60.9 | 68.4 | 72.7 | 62.1 | 91,397 |
| Black | 17.7 | 13.2 | 9.2 | 6.9 | 12.5 | 12,346 |
| Latino | 21.0 | 17.3 | 14.4 | 6.7 | 16.4 | 18,336 |
| Chinese | 1.3 | 1.0 | 1.2 | 2.1 | 1.2 | 1,793 |
| Filipino | 1.0 | 1.0 | 0.9 | 1.9 | 1.0 | 1,333 |
| Vietnamese | 0.5 | 0.8 | 0.4 | 1.5 | 0.6 | 798 |
| Korean | 0.5 | 0.5 | 0.6 | 1.5 | 0.6 | 811 |
| Other Asian | 2.1 | 1.9 | 1.7 | 4.4 | 2.1 | 2,607 |
| Native American | 1.0 | 0.6 | 0.6 | 0.3 | 0.7 | 1,093 |
| Pacific Islander | 0.2 | 0.2 | 0.2 | 0.1 | 0.2 | 216 |
| Mixed Race | 2.7 | 2.4 | 2.3 | 1.7 | 2.4 | 2,950 |
| Other Race | 0.2 | 0.3 | 0.2 | 0.2 | 0.2 | 261 |
| Total Percent | 100 | 100 | 100 | 100 | 100 | |
| Total N | 33,212 | 41,527 | 48,165 | 11,037 | 133,941 | |

Note: Data are from the 2018 American Community Survey (ACS). Percentages are calculated using weights provided by the ACS; sample sizes (N) are unweighted. Workers are considered to be at highest risk of interactions with external customers or the public if the average level of interactions with external customers or the public for their occupation in the O*NET data falls in the highest quartile of the occupations in the analysis. Occupational standing is defined as the percentage of ACS respondents reporting this occupation title who completed at least one year of college education. Occupational standing is then divided into quartiles. The 1st quartile represents the lowest quartile of occupational standing and the 4th quartile the highest quartile of occupational standing. The percentages in this table correspond to those displayed in panel 6 of Figs 2 and 4.

**Table S1.13: Racial/ethnic distribution of female frontline workers at highest risk of working indoors by quartile of occupational standing**

| Race/Ethnicity | Quartile of occupational standing | | | | | |
|---|---|---|---|---|---|---|
| | 1st Quartile | 2nd Quartile | 3rd Quartile | 4th Quartile | All in risk quartile | Total N |
| White | 51.6 | 62.9 | 61.7 | 71.0 | 67.0 | 67,563 |
| Black | 8.4 | 12.3 | 16.2 | 9.8 | 11.5 | 7,816 |
| Latino | 26.7 | 15.6 | 14.1 | 7.1 | 10.6 | 8,249 |
| Chinese | 3.2 | 1.3 | 0.8 | 1.5 | 1.4 | 1,327 |
| Filipino | 0.9 | 0.7 | 1.8 | 3.1 | 2.4 | 2,192 |
| Vietnamese | 2.4 | 1.6 | 0.3 | 0.8 | 0.8 | 724 |
| Korean | 1.8 | 0.8 | 0.3 | 0.9 | 0.8 | 691 |
| Other Asian | 3.0 | 2.1 | 1.4 | 3.4 | 2.7 | 2,321 |
| Native American | 0.3 | 0.5 | 0.8 | 0.3 | 0.5 | 520 |
| Pacific Islander | 0.1 | 0.1 | 0.1 | 0.1 | 0.1 | 81 |
| Mixed Race | 1.4 | 2.0 | 2.3 | 1.9 | 2.0 | 1,614 |
| Other Race | 0.1 | 0.2 | 0.3 | 0.2 | 0.2 | 156 |
| Total Percent | 100 | 100 | 100 | 100 | 100 | |
| Total N | 3,777 | 12,149 | 19,573 | 57,755 | 93,254 | |

Note: Data are from the 2018 American Community Survey (ACS). Percentages are calculated using weights provided by the ACS; sample sizes (N) are unweighted. Because working indoors rather than outdoors reflects high risk, we take a maximum of the O*NET values for two variables measuring working outdoors and designate workers whose occupations fall in the lowest quartile of this measure as having the highest risk of working indoors. Occupational standing is defined as the percentage of ACS respondents reporting this occupation title who completed at least one year of college education. Occupational standing is then divided into quartiles. The 1st quartile represents the lowest quartile of occupational standing and the 4th quartile the highest quartile of occupational standing. The percentages in this table correspond to those displayed in panel 7 of Figs 2 and 4.
