## Supplementary material for "Racial and ethnic differentials in COVID-19-related job exposures by occupational standing in the US": S2 Appendix

**S2 Appendix: Common high-risk occupations by occupational standing quartile**

*Updated April 5, 2021*

**Table S2.1: Common high-risk occupations held by males by occupational standing quartile**

| Occupational standing Quartile | Occupations at highest risk of exposure to infection | Percent |
|---|---|---|
| 1st quartile | janitors and building cleaners | 62.8 |
| | pipelayers, plumbers, pipefitters, and steamfitters | 18.4 |
| | maids and housekeeping cleaners | 6.7 |
| 2nd quartile | maintenance and repair workers, general | 21.9 |
| | bus and ambulance drivers and attendants | 14.0 |
| | personal care aides | 13.7 |
| | sheriffs, bailiffs, correctional officers, and jailers | 11.9 |
| | nursing, psychiatric and home health aides | 10.7 |
| | first-line supervisors of housekeeping and janitorial workers | 5.9 |
| | telecommunications line installers and repairers | 5.4 |
| 3rd quartile | police officers and detectives | 35.9 |
| | firefighters | 13.6 |
| | medical assistants and other healthcare support occupations, other | 7.3 |
| | emergency medical technicians and paramedics | 7.0 |
| | health diagnosing and treating practitioner support technicians | 6.7 |
| | licensed practical and licensed vocational nurses | 5.7 |
| 4th quartile | physicians and surgeons | 30.6 |
| | registered nurses | 21.7 |
| | pharmacists | 7.2 |
| | diagnostic related technologists and technicians | 6.8 |
| | dentists | 6.0 |
| | clinical laboratory technologists and technicians | 5.1 |
| | **Occupations at highest risk of physical proximity to others** | |
| 1st quartile | carpenters | 45.4 |
| | combined food preparation and serving workers, including fast food | 9.0 |
| | food preparation and serving related workers, other | 8.7 |
| | roofers | 8.5 |
| | butchers and other meat, poultry, and fish processing workers | 6.8 |
| | brickmasons, blockmasons, and stonemasons | 6.1 |
| 2nd quartile | first-line supervisors of construction trades and extraction workers | 17.4 |
| | waiters and waitresses | 15.9 |
| | taxi drivers and chauffeurs | 13.1 |
| | bus and ambulance drivers and attendants | 7.5 |

| | | |
|---|---|---|
| | industrial and refractory machinery mechanics | 7.4 |
| | personal care aides | 7.3 |
| | sheriffs, bailiffs, correctional officers, and jailers | 6.4 |
| | nursing, psychiatric, and home health aides | 5.7 |
| | first-line supervisors of food preparation and serving workers | 5.3 |
| 3rd quartile | firefighters | 17.7 |
| | recreation and fitness workers | 14.2 |
| | bartenders | 11.9 |
| | medical assistants and other healthcare support occupations, other | 9.5 |
| | emergency medical technicians and paramedics | 9.1 |
| | health diagnosing and treating practitioner support technicians | 8.7 |
| | licensed practical and licensed vocational nurses | 7.5 |
| | first-line supervisors of police and detectives | 6.1 |
| 4th quartile | physicians and surgeons | 32.4 |
| | registered nurses | 22.9 |
| | aircraft pilots and flight engineers | 9.8 |
| | diagnostic related technologists and technicians | 7.1 |
| | dentists | 6.3 |
| | **Occupations at highest risk of face-to-face discussions** | |
| 1st quartile | stock clerks and order fillers | 24.8 |
| | carpenters | 24.7 |
| 2nd quartile | retail salespersons | 42.7 |
| | first-line supervisors of construction trades and extraction workers | 19.1 |
| | maintenance and repair workers, general | 12.9 |
| | first-line supervisors of mechanics, installers, and repairers | 6.0 |
| | supervisors of transportation and material moving workers | 5.4 |
| 3rd quartile | first-line supervisors of sales workers | 78.2 |
| | insurance sales agents | 8.8 |
| 4th quartile | physicians and surgeons | 36.9 |
| | registered nurses | 26.1 |
| | pharmacists | 8.6 |
| | dentists | 7.2 |
| | **Occupations at highest risk of interactions with external customers or the public** | |
| 1st quartile | cashiers | 79.9 |
| | automotive and watercraft service attendants | 6.6 |
| | vehicle and mobile equipment mechanics, installers, and repairers, other | 6.3 |
| 2nd quartile | retail salespersons | 50.3 |
| | sheriffs, bailiffs, correctional officers, and jailers | 8.2 |

| | | |
|---|---|---|
| | first-line supervisors of mechanics, installers, and repairers | 7.1 |
| | first-line supervisors of food preparation and serving workers | 6.9 |
| | law enforcement workers, other | 5.5 |
| 3rd quartile | first-line supervisors of sales workers | 40.1 |
| | sales representatives, wholesale and manufacturing | 15.3 |
| | police officers and detectives | 11.9 |
| | sales representatives, services, all other | 7.4 |
| 4th quartile | pharmacists | 22.6 |
| | diagnostic related technologists and technicians | 21.3 |
| | dentists | 18.9 |
| | physical therapists | 12.3 |
| | respiratory therapists | 6.5 |
| | sales engineers | 5.7 |
| | veterinarians | 5.6 |
| | **Occupations at highest risk of working indoors** | |
| 1st quartile | bookbinders, printing machine operators, and job printers | 32.2 |
| | barbers | 24.8 |
| | sewing machine operators | 11.7 |
| | upholsterers | 6.3 |
| | extruding, forming, pressing, and compacting machine setters, operators, and tenders | 6.3 |
| 2nd quartile | bakers | 26.4 |
| | hairdressers, hairstylists, and cosmetologists | 22.0 |
| | gaming services workers | 18.0 |
| | medical, dental, and ophthalmic laboratory technicians | 11.9 |
| | telemarketers | 9.0 |
| | jewelers and precious stone and metal workers | 6.6 |
| 3rd quartile | bartenders | 44.8 |
| | licensed practical and licensed vocational nurses | 28.1 |
| | health technologists and technicians, other | 10.9 |
| 4th quartile | physicians and surgeons | 32.0 |
| | registered nurses | 22.6 |
| | securities, commodities, and financial services sales agents | 9.4 |
| | pharmacists | 7.5 |
| | diagnostic related technologists and technicians | 7.1 |
| | dentists | 6.3 |
| | clinical laboratory technologists and technicians | 5.3 |

Note: Occupational standing is defined as the percentage of ACS respondents reporting this occupation title who completed at least one year of college education. Occupational standing is then divided into quartiles. The 1st quartile represents the lowest quartile of occupational standing and the 4th quartile the highest quartile of occupational standing. Information on the five occupational risk factors for COVID-19 is obtained from O*NET; occupations in the top ($4^{th}$) quartile for each of these risk factors are considered to be at highest risk. Values are the percentage of workers employed in the particular combination of occupational standing quartile and risk factor quartile who report the listed job title. For brevity, only occupations exceeding 5% are displayed.

**Table S2.2: Common high-risk occupations held by females by occupational standing quartile**

| Occupational standing Quartile | Occupations at highest risk of exposure to infection | Percent |
|---|---|---|
| 1st Quartile | maids and housekeeping cleaners | 52.9 |
| | janitors and building cleaners | 35.8 |
| | food servers, nonrestaurant | 5.0 |
| 2nd Quartile | nursing, psychiatric, and home health aides | 29.0 |
| | childcare workers | 23.1 |
| | personal care aides | 22.7 |
| | hairdressers, hairstylists, and cosmetologists | 12.4 |
| 3rd Quartile | medical assistants and other healthcare support occupations, other | 26.0 |
| | licensed practical and licensed vocational nurses | 24.2 |
| | health diagnosing and treating practitioner support technicians | 17.8 |
| | dental assistants | 8.6 |
| 4th Quartile | registered nurses | 56.7 |
| | physicians and surgeons | 6.2 |
| | **Occupations at highest risk of physical proximity to others** | |
| 1st Quartile | combined food preparation and serving workers, including fast food | 37.2 |
| | host and hostesses, restaurant, lounge, and coffee shop | 29.0 |
| | food preparation and serving related workers, other | 13.9 |
| | butchers and other meat, poultry, and fish processing workers | 6.2 |
| 2nd Quartile | nursing, psychiatric, and home health aides | 21.0 |
| | waiters and waitresses | 20.3 |
| | childcare workers | 16.7 |
| | personal care aides | 16.5 |
| | hairdressers, hairstylists, and cosmetologists | 9.0 |
| 3rd Quartile | medical assistants and other healthcare support occupations, other | 22.9 |
| | licensed practical and licensed vocational nurses | 21.4 |
| | health diagnosing and treating practitioner support technicians | 15.7 |
| | recreation and fitness workers | 9.9 |
| | dental assistants | 7.6 |
| | bartenders | 7.5 |
| 4th Quartile | registered nurses | 63.7 |
| | physicians and surgeons | 6.9 |
| | diagnostic related technologists and technicians | 5.5 |
| | **Occupations at highest risk of face-to-face discussions** | |
| 1st Quartile | stock clerks and order fillers | 51.7 |
| | laborers and freight, stock, and material movers, hand | 42.4 |
| 2nd Quartile | retail salespersons | 60.1 |

| | | |
|---|---|---|
| | hairdressers, hairstylists, and cosmetologists | 23.7 |
| | nonfarm animal caretakers | 7.6 |
| 3rd Quartile | first-line supervisors of sales workers | 59.4 |
| | licensed practical and licensed vocational nurses | 25.2 |
| | insurance sales agents | 8.2 |
| 4th Quartile | registered nurses | 66.8 |
| | physicians and surgeons | 7.3 |
| | **Occupations at highest risk of interactions with external customers or the public** | |
| 1st Quartile | cashiers | 97.9 |
| 2nd Quartile | retail salespersons | 53.3 |
| | hairdressers, hairstylists, and cosmetologists | 21.1 |
| | first-line supervisors of food preparation and serving workers | 9.4 |
| | law enforcement workers, other | 5.3 |
| 3rd Quartile | first-line supervisors of sales workers | 45.0 |
| | sales representatives, wholesale and manufacturing | 9.3 |
| | dental assistants | 6.8 |
| | bartenders | 6.6 |
| | insurance sales agents | 6.2 |
| | door-to-door sales workers, news and street vendors, and related workers | 6.2 |
| | sales representatives, services, all other | 5.1 |
| 4th Quartile | diagnostic related technologists and technicians | 28.8 |
| | pharmacists | 19.6 |
| | physical therapists | 17.9 |
| | dieticians and nutritionists | 10.5 |
| | respiratory therapists | 7.7 |
| | dentists | 5.8 |
| | veterinarians | 5.6 |
| | **Occupations at highest risk of working indoors** | |
| 1st Quartile | sewing machine operators | 43.6 |
| | tailors, dressmakers, and sewers | 19.1 |
| | bookbinders, printing machine operators, and job printers | 15.5 |
| | barbers | 8.6 |
| 2nd Quartile | hairdressers, hairstylists, and cosmetologists | 71.0 |
| | bakers | 13.7 |
| 3rd Quartile | licensed practical and licensed vocational nurses | 46.2 |
| | dental assistants | 16.4 |
| | bartenders | 16.1 |
| | medical records and health information technicians | 9.4 |
| | health technologists and technicians, other | 5.5 |
| 4th Quartile | registered nurses | 64.3 |

| | |
|---|---|
| physicians and surgeons | 7.0 |
| diagnostic related technologists and technicians | 5.5 |
| clinical laboratory technologists and technicians | 5.2 |

Note: Occupational standing is defined as the percentage of ACS respondents reporting this occupation title who completed at least one year of college education. Occupational standing is then divided into quartiles. The 1st quartile represents the lowest quartile of occupational standing and the 4th quartile the highest quartile of occupational standing. Information on the five occupational risk factors for COVID-19 is obtained from O*NET; occupations in the top ($4^{th}$) quartile for each of these risk factors are considered to be at highest risk. Values are the percentage of workers employed in the particular combination of occupational standing quartile and risk factor quartile who report the listed job title. For brevity, only occupations exceeding 5% are displayed.
